# COVID-19 with early neurological and cardiac thromboembolic phenomena—timeline of incidence and clinical features

**DOI:** 10.1101/2021.03.15.21253619

**Authors:** Uma Sundar, Sanah Merchant, Meera Shah, Amita Mukhopadhyay, Shaonak Kolte, Pramod Darole, Sharvari Mahajan, Ashank Bansal, Satish Gosavi, Dnaneshwar Asole, Niteen D Karnik, Ajay Mahajan, Anagha Joshi

## Abstract

**Background:** At our tertiary care public hospital, we saw COVID-19 presenting with thromboembolic phenomena, indicating a possible early thrombo-inflammatory pathology.

**Objectives:** We documented patients with cardiac and neurological thromboembolic phenomena as a primary presentation of COVID-19, and compared a subset of COVID associated strokes against COVID-19 patients without thrombotic manifestations.

**Methods:** We included all COVID-Stroke and COVID-ACS (COVID-19, with ischemic arterial stroke/Acute Coronary Syndrome presenting prior to/simultaneous with/within 72 hours of systemic/respiratory COVID manifestations) admitted from April to November 2020. In the nested case control analysis, we used unpaired T-test and chi-square test to study differences between COVID-Strokes (case group) and non-thrombotic COVID controls.

**Results and Conclusions:** We noted 68 strokes and 122 ACS associated with COVID-19. ACS peaked in May-June, while stroke admissions peaked later in September-October, possibly because severe strokes may have expired at home during the lockdown.

In the case-control analysis, cases (n=43; 12F:31M; mean age 51.5 years) had significantly higher D-Dimer values than controls (n=50; 9F:41M; mean age 51.6 years). Mortality was significantly higher in cases (51.2% vs. 26.0%; p = 0.018). We noted 7.5 times higher mortality in cases versus controls even among patients needing minimal oxygen support. Imaging in 37 patients showed both anterior and posterior circulation territories affected in seven, with almost half of Carotid territory strokes being large hemispherical strokes. Additionally, CT/MRI angiography in 28 strokes showed large vessel occlusions in 19 patients. Death in cases thus probably occurred before progression to intense respiratory support, due to severe central nervous system insult.

Binary logistic regression analysis showed respiratory support intensity to be the sole independent predictor of mortality among cases. Respiratory distress could have been due to COVID-19 lung infection or aspiration pneumonia resulting from obtunded sensorium. In controls, mortality was predicted by increasing age, female sex, and respiratory support intensity.

## Introduction

COVID-19 is caused by the SARS-CoV-2 virus, a member of the Coronaviridae family. There are numerous reports of patients with COVID-19 presenting with both arterial (stroke, myocardial infarction) and venous thrombosis (deep vein thrombosis, pulmonary thromboembolism, cerebral venous sinus thrombosis).(1–10) Accelerated thrombogenesis in COVID-19 is postulated to be due to Virchow’s triad, resulting in endothelial dysfunction, abnormal flow and a hypercoagulable state.(11–13) At our tertiary care public hospital, we saw COVID-19 presenting with thromboembolic phenomena, where the respiratory involvement was either very mild and simultaneous, or occurred only later during hospitalization, indicating a possible early thrombo-inflammatory pathology, in a subsection of patients. Hence, we documented patients with cardiac and neurological thromboembolic phenomena as a primary presentation of COVID-19. Further, we compared clinical and inflammatory markers in a subset of these patients with markers in patients without thrombotic manifestations of the disease during hospitalization.

## Aims

We aimed to document the incidence of acute thromboembolism, namely, ischemic stroke and acute coronary syndrome (ACS) as a presentation of COVID-19. We also aimed to compare clinical and inflammatory markers in a subgroup of patients of COVID-19 with acute ischemic stroke at presentation or within the first 72 hours of other signs and symptoms of COVID-19, with the parameters in patients presenting with respiratory onset disease. This latter group had neither cardiac nor neurological ischemic pathology during their hospitalization.

## Methodology

Our study center is a 1400 bedded, tertiary care, municipal public hospital in Mumbai, India. We did a retrospective observational serial recruitment of all cases satisfying the inclusion criteria, followed by a nested case control analysis of a subset of the larger cohort of COVID-19 patients.

### Operational definitions

#### Ischemic stroke

Patients who presented to the hospital with ischemic arterial stroke, without systemic or respiratory signs and symptoms of COVID-19 at admission, or stroke developing within 72 hours of systemic or respiratory signs and symptoms of COVID-19, were included from a study period of April to November 2020. Any of the symptoms comprising fever, myalgia, throat pain, dry cough, tachypnoea, loose motions, and an oxygen saturation under 95% on room air at admission were considered as indicative of presence of systemic and respiratory signs of COVID-19.

#### Acute coronary syndrome

Patients who presented to the hospital with ACS {ST segment elevation myocardial infarction (STEMI) or Non-ST segment elevation myocardial infarction (NSTEMI), or Unstable angina, diagnosed as per standard criteria} without systemic or respiratory signs and symptoms of COVID-19 at admission, or ACS developing within 72 hours of systemic or respiratory signs and symptoms of COVID-19, were included in a similar manner.

Patients with pulmonary or other systemic and peripheral thromboembolism and patients with cerebral venous sinus thrombosis (CVST) were not included in the detailed analysis in this study, unless associated with stroke or ACS, as a separate registry is being maintained for them.

### Inclusion in case group and control group

We compared clinical and inflammatory markers inpatients of COVID-19 with thromboembolic phenomena (acute ischemic stroke) at presentation or within the first 72 hours of other signs and symptoms of COVID-19, with the parameters in patients presenting with respiratory onset disease. The latter group continued to be free of cardiac and neurological ischemic pathology till discharge. We selected the group with acute ischemic stroke as the case group, as complete data was available for the patients with COVID-19 associated stroke. Thus, we analyzed detailed data for 43 COVID-19 cases with associated stroke, and 50 controls with COVID-19 and no associated thromboembolic events.

Controls were included in a number similar to the cases. The selection of controls from any given month of the study duration (April to November 2020), corresponded to the proportion of COVID strokes to total COVID-19 cases, for that month. As strokes were highest in proportion to total COVID-19 cases in September and October 2020, the maximum number of controls were taken from that period. Selection of individual cases was by using random computer-generated numbers.

We evaluated and compared demographic features (age, gender), clinical features (comorbidities, oxygen saturation at admission and modality of maximum oxygen support required during hospitalization), laboratory parameters {inflammatory markers such as C Reactive Protein (CRP) and D-Dimer, creatinine level, platelet count}, mortality and predictors of mortality in the case group versus the control group. Modalities of respiratory support included, in order of increasing intensity, nasal cannula, venturi mask, non-rebreathing bag and mask (NRBM), high flow nasal cannula (HFNC), non-invasive ventilation (NIV), and intubation and mechanical ventilation. In the correlation of in-hospital mortality with requirement for respiratory support, we divided the patients into two groups. The low flow oxygen support subgroup included patients on room air, nasal cannula and venturi mask, and the high flow oxygen support subgroup included patients on NRBM, HFNC, NIV and intubation with mechanical ventilation.

We used Microsoft Excel (2010) to tabulate and clean the raw data, and to generate pivot tables and graphs. We imported the excel datasheet into IBM SPSS version 20.0 and performed univariate and multivariate analyses. We divided the study set into subgroups according to relevant characteristics, and used the unpaired samples T-test and chi-square test to study differences in key variables. We assumed unequal variances in reporting T tests. We calculated 95% confidence intervals for mean differences and odds ratios, and considered a p value below 0.05 to be statistically significant only if the confidence intervals for the concerned statistic were congruent. We performed binary logistic regression to determine the key predictors of in-hospital mortality. We entered the following variables as independent predictorsfor both case and control groups: age, sex, number of comorbidities and respiratory distress level (the last coded on an ascending scale reflecting the intensity of mode of supplementary oxygen delivery, where 0 represented no support required, and 6 represented invasive ventilation). CRP and D-Dimer were not added as predictors to this model, as laboratory reports for both markers were available for only 20 cases. In the initial days of the pandemic, logistic pressures on the hospital made it difficult to procure reports for all patients.

#### Ethical considerations

We obtained scientific and ethical approval for the study from the Human Research Institutional Ethics Committee, Lokmanya Tilak Municipal Medical College and General Hospital, Sion, Mumbai.

#### Data availability statement

The data used in our analysis will be made available to researchers upon reasonable request to the corresponding author.

## Results

Figure 1 shows that during the period of April-November 2020, there were 4,069 COVID-19 cases admitted in our hospital. There were 68 cases of stroke associated with COVID-19 and 122 cases of ACS associated with COVID-19. The peak of total COVID-19 cases and cardiac cases was in May and June 2020.The peak of stroke cases was in September and October 2020. Among the 68 patients with stroke, 49 had ischemic stroke. Of these 49 patients, 43 satisfied inclusion criteria for early presentation as ischemic stroke as per our operational definitions. The case records of these 43 patients (case group) were analyzed in detail. Close to half of the 43 patients in the case group (21; 48.9%) belonged to the 41-60 years age group. Among them, 11 were aged 51-60 years, and 10 were 41-50 years of age. Whereas out of 50 controls, 13 patients (26.0%) were aged 61-70 years, followed by 11 each (22.0%) in the 31-40 years and 51-60 years age groups, respectively.

**Figure 1.**
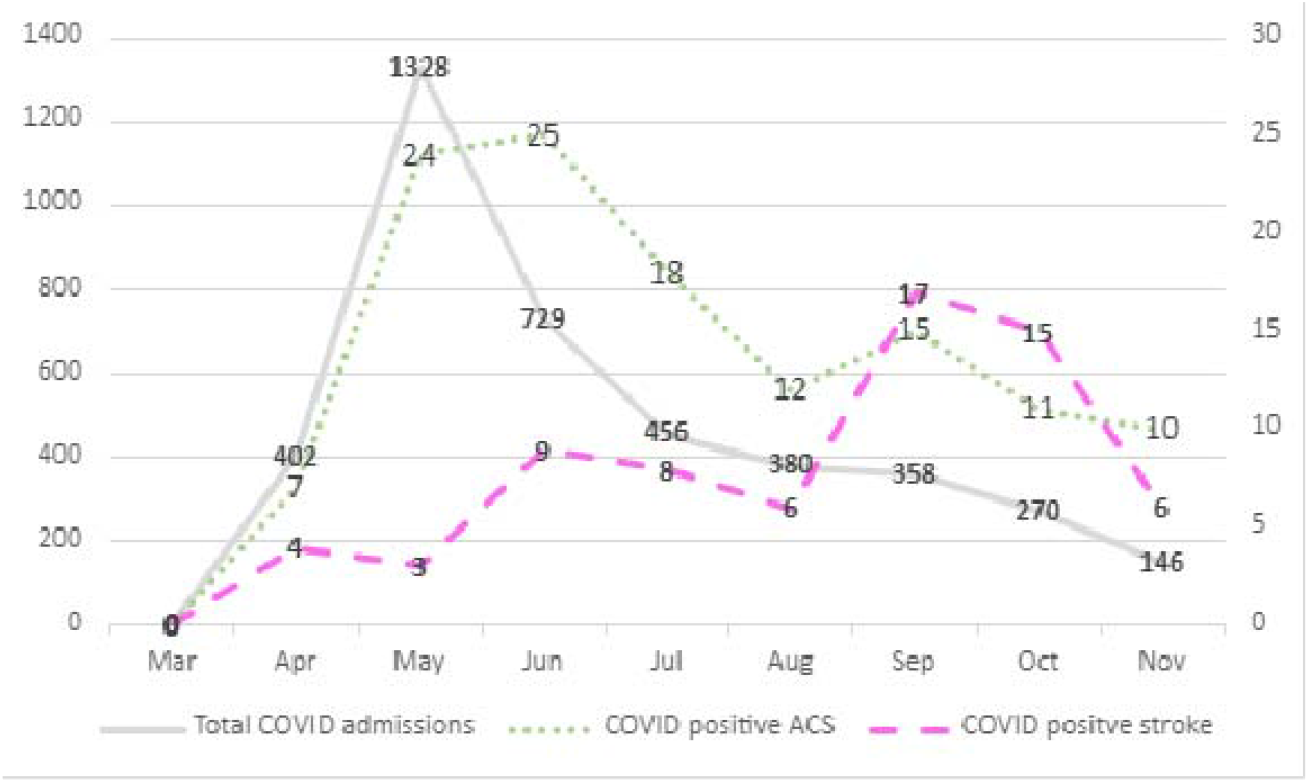
Time trend of COVID-19cases, strokes, and acute coronary syndromes ACS: Acute Coronary Syndrome

The sex distribution of cases and controls was as follows: 12 (27.9%) women and 31 (72.1%) men in the case group, versus 9 (18.0%) women and 41 (82.0%) men in the control group.

There was no significant difference in the age or sex distribution of case and control groups.

Among the cases, 14 of 43 patients (32.6%) were free of comorbidities, compared to 22 of 50 controls (44.0%). Among the cases there were 9 patients with hypertension (HTN), 4 with diabetes mellitus (DM), and 12 who had both comorbidities. Among the controls, 6 patients had HTN, 11 had DM, and 9 had both. These differences were not statistically significant.

Among the 43 stroke cases, we noted 40 patients with pure arterial infarcts, one with arterial infarct associated with CVST, and two patients with bithalamic infarcts, possibly due to blockage of artery of Percheron or vein of Galen. As CT angiography and venography were not available for these two patients, they were classified as of indeterminate (arterial/venous) etiology. CT scans pinpointed the vascular territory affected in 37 of 41 patients (40 pure arterial infarcts and one infarct+CVST) with arterial infarcts. Of these,26 (70.3% of 37) had infarcts in the carotid territory, four (10.8% of 37) in the vertebrobasilar territory, and seven patients (18.9% of 37) had infarcts in both territories. Additionally, four patients were clinically diagnosed as carotid territory infarct, although their initial CT scans revealed no acute changes (repeat CTs were not done either due to logistic constraints or clinical deterioration). Among the 33 carotid ischemic strokes with CT scans available (comprising carotid territory strokes and carotid component of mixed territory strokes), 15 were classified as large hemispherical, 10 were subcortical and 8 were pure cortical infarcts.

Among the 43 stroke patients, 28 (65.1%) had a CT angiography or MRI angiography done. Large vessel occlusion (LVO) was present in 19/28 (67.8%) patients.

As seen in Table 1, cases had significantly lower average levels of respiratory support requirement and significantly higher D-Dimer values than controls.

**Table 1.** Comparison of means for selected variables in the case and control groups.

| Variables |  |  |  | Mean<br>Difference | 95% Confidence<br>Interval of the<br>Difference |  | P<br>value |
| --- | --- | --- | --- | --- | --- | --- | --- |
|  |  | N | Mean |  | Lower | Upper |  |
| Age | Case | 43 | 51.51 | -0.11 | -6.29 | 6.08 | 0.972 |
|  | Control | 50 | 51.62 |  |  |  |  |
| No. of<br>Comorbidities | Case | 43 | 1.00 | 0.20 | -0.15 | 0.55 | 0.255 |
|  | Control | 50 | .80 |  |  |  |  |
| SpO<sub>2</sub> on Admission | Case | 43 | 89.12 | 2.70 | -3.05 | 8.45 | 0.354 |
|  | Control | 50 | 86.42 |  |  |  |  |
| Respiratory<br>Distress Level | Case | 43 | 1.14 | -1.58 | -2.27 | -0.89 | 0.000 |
|  | Control | 50 | 2.72 |  |  |  |  |
| Platelet count/μL | Case | 39 | 214925.64 | 3160.42 | -55915.90 | 62236.75 | 0.914 |
|  | Control | 23 | 211765.22 |  |  |  |  |
| S. Creatinine<br>mg/dL | Case | 41 | 1.24 | -0.01 | -0.31 | 0.29 | 0.936 |
|  | Control | 50 | 1.26 |  |  |  |  |
| S. Creatinine<br>μmol/L | Case | 41 | 109.99 | -1.07 | -27.69 | 25.55 | 0.936 |
|  | Control | 50 | 111.06 |  |  |  |  |
| CRP mg/L | Case | 20 | 59.63 | -20.73 | -53.77 | 12.30 | 0.212 |
|  | Control | 50 | 80.36 |  |  |  |  |
| D-dimer ng/mL | Case | 20 | 2975.44 | 1472.02 | 110.72 | 2833.32 | 0.035 |
|  | Control | 50 | 1503.42 |  |  |  |  |
| D-dimer nmol/L | Case | 20 | 32.59 | 16.12 | 1.21 | 31.03 | 0.035 |
|  | Control | 50 | 16.47 |  |  |  |  |

SpO2: percentage saturation of oxygen
CRP: C-Reactive protein

Comparing maximum respiratory support requirement in cases versus controls, we noted that 32/50 (64.0%) of the controls required intensive modes of respiratory support (NRBM, HFNC, NIV, Intubation) as compared to 10/43 (23.26%) of the cases. This difference was statistically significant (Χ^2^ = 2.022, df 6, p = 0.001).

Mortality was 51.2% in the case group compared with26.0% in the control group, being significantly higher in cases compared to controls (Χ^2^ =6.237, df 1, p = 0.018; OR 2.98, 95CI_OR_ 1.25–7.11).

Table 2 shows that patients requiring higher modalities of oxygen support (NRBM, HFNC, NIV, invasive ventilation) had higher mortality in both cases and controls. We observed that in both low flow oxygen and high flow oxygen subgroups, cases had significantly higher mortality than controls. However, the difference in mortality between cases and controls was striking in the low flow subgroup. In the latter, despite maintaining satisfactory oxygen saturation on room air or with minimal oxygen support, the mortality was 7.5 times higher in COVID stroke patients as compared to controls (Low flow: 42.4% vs 5.6%, Χ^2^= 7.626, df 1, p = 0.006; OR 12.53, 95CI_OR_ 1.49–105.58; High flow: 80.0% vs 37.5%, Χ^2^= 5.517, df 1, p = 0.030; OR 6.67, 95CI_OR_ 1.21–36.74).

**Table 2.** Maximum respiratory support requirement versus mortality in case and control groups

|  | Case |  |  |  | Control |  |  |  |
| --- | --- | --- | --- | --- | --- | --- | --- | --- |
|  | Died |  | Survived |  | Died |  | Survived |  |
|  | Count | % | Count | % | Count | % | Count | % |
| Room air | 5 | 25.0 | 15 | 75.0 | 0 | 0.0 | 8 | 100.0 |
| Nasal cannula | 7 | 70.0 | 3 | 30.0 | 1 | 10.0 | 9 | 90.0 |
| Venturi | 2 | 66.7 | 1 | 33.3 | 0 | 0.0 | 0 | 0.0 |
| NRBM | 6 | 75.0 | 2 | 25.0 | 2 | 10.5 | 17 | 89.5 |
| HFNC | 1 | 100.0 | 0 | 0.0 | 2 | 50.0 | 2 | 50.0 |
| NIV | 1 | 100.0 | 0 | 0.0 | 0 | 0.0 | 1 | 100.0 |
| Intubation | 0 | 0.0 | 0 | 0.0 | 8 | 100.0 | 0 | 0.0 |
| Low flow group | 14 | 42.4 | 19 | 57.6 | 1 | 5.6 | 17 | 94.4 |
| High flow group | 8 | 80.0 | 2 | 20.0 | 12 | 37.5 | 20 | 62.5 |

Low flow: Room air, Nasal canula and Venturi modes
High flow: NRBM, HFNC, NIV and Intubation modes

Accurate dates of admission and discharge/death were available for 26 of 43 cases, and all 50 controls. Mean duration of hospitalization for cases was 13.25 days for those who survived and 9.36 days for those who died. For controls it was 12.57 days for those who survived and 11.54 days for those who died.

Table 3 shows the results of binary logistic regression analysis to determine predictors of mortality in the case group and control group. The level of respiratory support requirement was the sole independent predictor of mortality among the cases. In controls, mortality was predicted by increasing age and female sex in addition to respiratory support requirement.

**Table 3.** Regression analysis for predictors of mortality in cases and controls

| CASE/CONTROL |  | B | Exp(B) | 95% C.I. for EXP(B) |  | Sig. |
| --- | --- | --- | --- | --- | --- | --- |
|  |  |  |  | Lower | Upper |  |
| Case | Age | -.038 | .963 | .907 | 1.022 | .211 |
|  | Sex(F) | .896 | 2.450 | .472 | 12.731 | .287 |
|  | No. of comorbidities | .558 | 1.747 | .663 | 4.605 | .259 |
|  | Respiratory support requirement | 1.070 | 2.915 | 1.383 | 6.143 | .005 |
|  | Constant | .065 | 1.067 |  |  | .958 |
| Control | Age | .240 | 1.271 | 1.030 | 1.569 | .026 |
|  | Sex(F) | 5.823 | 337.932 | 1.984 | 57559.548 | .026 |
|  | No. of comorbidities | -2.406 | .090 | .007 | 1.235 | .072 |
|  | Respiratory support requirement | 2.700 | 14.877 | 1.917 | 115.460 | .010 |
|  | Constant | -23.396 | .000 |  |  | .012 |

## Discussion

During the COVID-19 pandemic, it was recognized early on that the presence of thrombosis in multiple organ systems points to thromboembolism being an integral component in the pathogenesis of this novel disease.(10,11,14,15)

During the months of May and June 2020, we noted many patients presenting with thromboembolic phenomena as an early manifestation of COVID-19, or developing these complications during admission. Hence, the first part of our study comprised documentation of acute cerebral and cardiac thromboembolic events during admission for COVID-19 infection. Acute neurovascular and cardiac events associated with COVID-19 at our centre included 68 cases of stroke and 122 cases of ACS during the period of April-November 2020. Of these 68 strokes, 49 had ischemic strokes. Of these 49 COVID-19 associated ischemic strokes, 22 (51.2%) presented with only stroke and no respiratory signs and symptoms of COVID-19 (data in press). This possible early thromboembolic effect of COVID-19 on the neurovascular system, prompted us to investigate and study patients with ischemic stroke as a primary presentation of COVID-19, in the second part of our study. The occurrence of thromboembolic phenomenon early in the course of COVID-19 disease has also been observed in other studies by Pillai et al,(14) Stefanini et al(16) and Mao et al.(17)

43 patients satisfied our inclusion criteria for this study viz., ischemic stroke within 72 hours of onset of first symptoms of COVID-19. As complete data was available for the patients with stroke, we compared characteristics of stroke patients with corresponding findings in patients without thromboembolic manifestations of the disease. A detailed similar analysis of the patients with ACS was not possible, due to lack of complete data.

Figure 1 shows the timeline of admission of all COVID-19 cases, COVID-19 cases with ACS and COVID-19 cases with stroke, under medicine and cardiology departments, at our centre. Whereas the peak of cardiac cases was in May and June 2020, the peak of stroke cases was in September and October 2020, at which time stroke and ACS incidence were approximately the same. As both ACS and stroke are thrombo-inflammatory manifestations of the virus, it is surprising that these two events have shown different peaks (Figure 1). ACS incidence showed a gradual decline after June whereas stroke incidence gradually rose to peak in September-October. In September and October 2020, the number of COVID-19 admissions (only patients with higher category oxygen requirement, beyond nasal cannula at 4 litres/minute, or those with systemic complications, are admitted at our centre) had fallen steeply, with ACS cases also showing a gradual decline. It is possible that patients with severe strokes may have expired at home due to difficulties of travel in a severely disabled state during the lockdown in peak months. Hence, the rise in stroke admissions later may have been due to better facilities of transportation, as the lockdown eased in the city in the later months.

Among the 43 stroke cases, CT confirmation of infarct and arterial territory could be made in 37 patients. Four patients were diagnosed clinically as carotid territory stroke, and the lack of hemorrhage on the CT classified them as possible early infarcts. Due to the difficulties inherent in and peculiar to the COVID-19 situation in a large public hospital, a repeat imaging study could not be done in these patients. However, it was clear that in those with imaging confirmation, the infarcts were either large or multiple, or in eloquent areas; 18.9% being in both anterior and posterior circulation territories, 10.8% in vertebrobasilar territory, and even among the carotid territory strokes, almost half being large hemispherical strokes. Additionally, 67.8% of those who had a CT angiography done, showed evidence of LVO, portending the possibility of enlargement of infarct area.

CRP levels, while being over 10 times the upper limit of normal level in both groups, showed no significant difference between the two groups (Table 1). This was not surprising as both the groups fell into the moderate to severe infection categories, in terms of respiratory and other systemic involvement. While both cases and controls had high levels of D-Dimer (more than thrice the upper limit of normal value), the case group had a significantly higher D-Dimer level as compared to the control group. A deranged coagulation function, including elevated D-Dimer, has been demonstrated to lead to disease progression of COVID-19.(18,19) Several studies have reported elevated levels of CRPs and D-Dimer in patients with COVID-19.(3,4,17,20,21) Additionally, COVID-19 associated inflammation resulting in a hypercoagulable state has been well documented.(21,22)

Our study showed that the control group required a higher level of respiratory support care than the case group, the number of patients requiring NRBM, HFNC or intubation being 32 (64%) in control group, as compared to 10 (23.26%) in case group. This could be related to two factors: the higher survival rate in the control group (mortality in case and control groups being 51.2% and 26.0% respectively), and longer mean duration of survival in the control group (the patients in the case group who died in hospital, survived for an average of 9.3 days as compared to 11.5 days in the patients who expired in the control group).

In the overall ‘low flow oxygen support group,’ the 7 times higher death rate in the case group as compared to the control group, was striking. Death in the case group probably occurred before progression to higher levels of respiratory support, due to severe central nervous system insult, without having severe respiratory involvement. The phenomenon of increased risk of thrombus formation leading to stroke (postulated to be due to viral involvement of the endothelium), in the absence of severe respiratory disease, has been previously documented.(15) Contrarily, other authors have posited that the incidence of stroke in COVID-19 may be related to severity of infection.(17,21)

In this study, although higher modes of respiratory support were less frequent in the case group, respiratory distress level requiring higher modality of support predicted mortality within the group. The respiratory distress could have been due to COVID-19 lung infection or due to aspiration pneumonia resulting from obtunded sensorium in stroke.

The prediction of mortality by increasing age, as seen in the control group in our study, has been well documented by various authors.(23,24) However, most studies have shown that men have a higher risk of COVID-19 related death than women,(23,25) contrary to our findings in the control group. Our sample size for the control group was small, and this limitation could have led to this finding.

A major limitation of our study is the lack of documentation of clinical and subclinical pulmonary thromboembolic phenomena in the case and control groups. This was highly probable, given the thrombo-inflammatory milieu, and the concomitantly existing cerebral thrombosis. It is possible that this was more common in the case group and could have contributed to the mortality. We did not take pulmonary thromboembolism into account in our analysis, as complete data on CT pulmonary angiography was not available in many patients.

In conclusion, our comparison of a thromboembolic cerebral presentation of COVID-19 with a clinically non-thromboembolic presentation of COVID-19, demonstrated higher D-Dimer levels, and a higher mortality in the absence of prominent respiratory compromise, in the former group. The higher mortality was possibly due to the severity of stroke and presence of proximal LVOs. We could not compare mortality in COVID-19 associated strokes with a cohort of non-COVID associated strokes in the same period, as our regular admissions were severely limited during the pandemic. A meta-analysis of stroke in COVID-19 has highlighted that the mean mortality rate among stroke patients with COVID-19 infection was 46.7% compared to only 8.7% among those without COVID-19 infection.(26)

## Supporting information

Strobe Case Control Checklist

Ethical Approval Document

