## Supplementary figures and images for "COVID-19 with early neurological and cardiac thromboembolic phenomena—timeline of incidence and clinical features"

### Ethical Approval Document

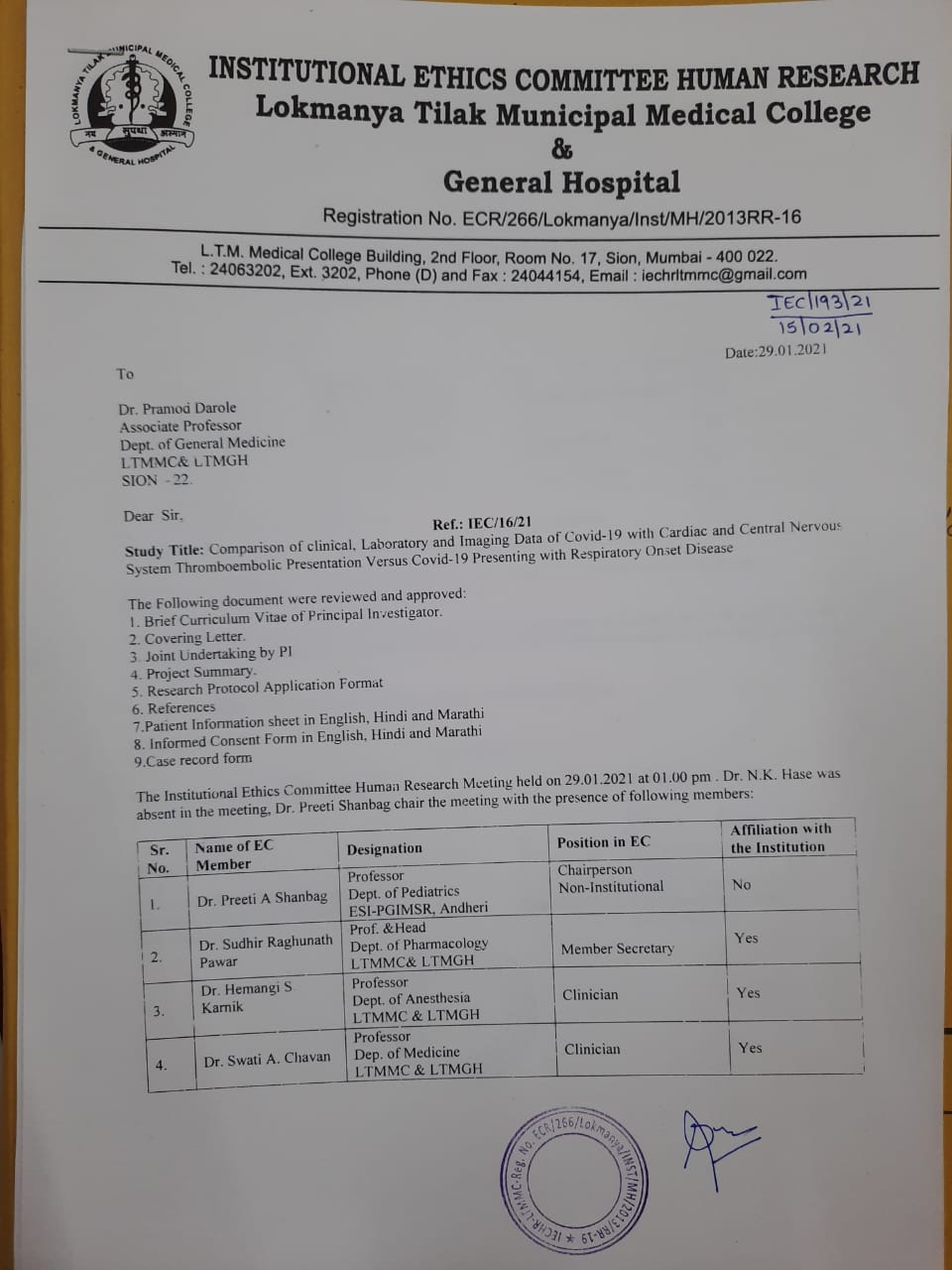

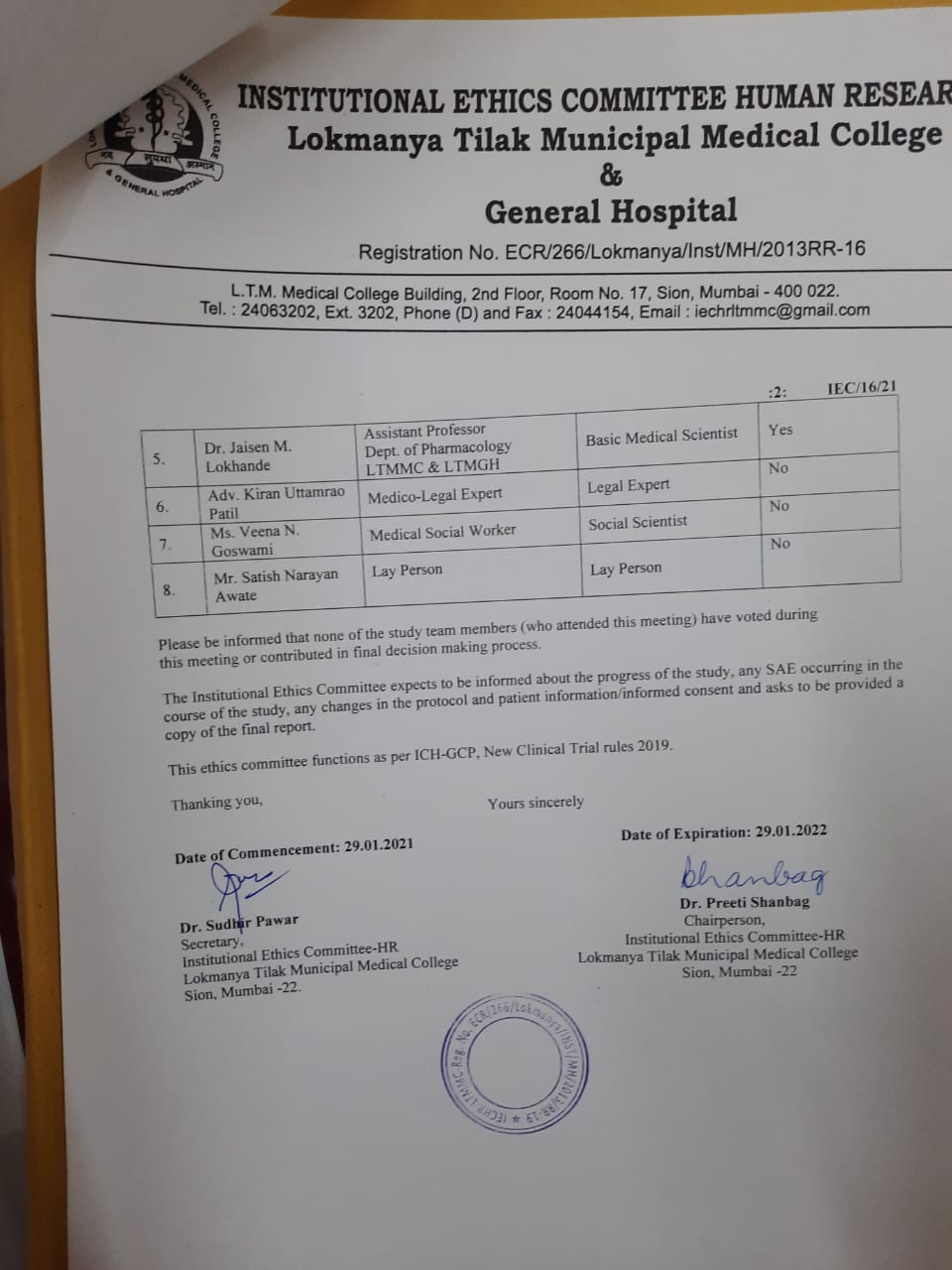
